# *Pseudomonas aeruginosa* in bronchiectasis: unmasking population diversity and antimicrobial resistance in the Australian context

**DOI:** 10.64898/2026.11.13.26367559

**Authors:** Kellie R. Strickland, Timothy Baird, Lucy D. Burr, Emily M. Dunn, Miriana L. Littlejohn, Jasmine L. Davies, Julia J. Bashford, Andrew Burke, Ieuan Evans, Jacob Christensen, Joshua Brock, Louise McIntosh, Erin P. Price, Derek S. Sarovich

**Affiliations:** Sunshine Coast Health Institute, Birtinya, Queensland, Australia; Centre for Bioinnovation, University of the Sunshine Coast, Sippy Downs, Queensland, Australia; School of Science, Technology and Engineering, University of the Sunshine Coast, Sippy Downs, Queensland, Australia; Department of Respiratory Medicine, Sunshine Coast University Hospital, Birtinya, Queensland, Australia; School of Medicine and Dentistry, Griffith University, Birtinya, Queensland, Australia; Mater Research Institute, University of Queensland, South Brisbane, Queensland, Australia; Department of Infectious Disease, The Prince Charles Hospital, Chermside, Queensland, Australia; Faculty of Health, Medicine and Behavioural Science, University of Queensland, St Lucia, Queensland, Australia; Department of Thoracic Medicine, The Prince Charles Hospital, Chermside, Queensland, Australia

**Keywords:** Bronchiectasis, antimicrobial resistance, whole-genome sequencing, comparative genomics, *Pseudomonas aeruginosa*, colistin, AMR prediction, phylogenomics

## Abstract

**Background:** *Pseudomonas aeruginosa* infections in bronchiectasis airways result in poorer lung function, advanced lung lobe involvement, worsened life quality, and higher hospitalisation rates. Despite high prevalence, little is known about *P. aeruginosa* population genomics, antimicrobial resistance (AMR) rates, persistence, and transmission dynamics in Australians with bronchiectasis.

**Methods:** *P. aeruginosa* (*n=*137) isolated between 2009 and 2025 from 59 Australians with bronchiectasis underwent antimicrobial susceptibility testing against 11 antipseudomonal drugs, whole-genome sequencing, comparative genomics, and *in silico* AMR determinant identification. Data were integrated with 4,575 publicly available bronchiectasis-derived *P. aeruginosa* genomes to determine global phylogenomic and AMR determinant patterns.

**Results:** High AMR rates were identified in the Australian isolates, with 75.9% resistant to at least one antimicrobial, 19.7% multidrug-resistant, and 9.5% extremely drug-resistant. AMR rates were highest (64.2%) for ciprofloxacin and cefepime (44.5%), and lowest for piperacillin/tazobactam (8.8%) and colistin (0.7%). Phylogenomics uncovered several instances of within-host evolution, hypermutation, persistence, and multi-lineage infections. Of the fifty-seven identified multilocus sequence types (STs), seven (12.3%) were novel, five were high-risk international clones (ST274 [6.6%], ST395 [0.7%], ST244 [0.7%], ST277 [0.7%], ST298 [0.7%]) and three were common global clones (ST253 [5.1%], ST155 [3.6%], ST260 [0.7%]). No high-risk AMR-conferring mobile genetic elements were seen, although numerous chromosomal AMR determinants were identified. AMR determinant prevalence was consistent with the global dataset, with no evidence of geographic clustering.

**Conclusions:** Our study provides critical insights into *P. aeruginosa* diversity in Australians with bronchiectasis. High AMR rates and chronic colonisation highlight the need to improve diagnostics, antimicrobial stewardship measures, and infection control practices in this neglected clinical cohort.

## INTRODUCTION

Antimicrobial resistance (AMR) is a leading threat to global health, with six priority pathogens, known collectively as the ESKAPE pathogens (***<u>E</u>****nterococcus faecium, **<u>S</u>**taphylococcus aureus, **<u>K</u>**lebsiella pneumoniae, **<u>A</u>**cinetobacter baumannii, **<u>P</u>**seudomonas aeruginosa* and ***<u>E</u>**nterobacter spp.)*, representing the greatest AMR burden (1,2). The environmentally ubiquitous ESKAPE pathogen, *Pseudomonas aeruginosa*, poses a significant concern across medicine, agriculture, and veterinary practice due its high virulence, capacity for extensive drug resistance (XDR), and association with poor clinical outcomes (3–5). *P. aeruginosa* is particularly burdensome for people with chronic respiratory conditions like cystic fibrosis (CF) and bronchiectasis. Bronchiectasis is characterised by permanent bronchial dilatation and distortion, resulting from a vicious cycle of infection, inflammation, impaired mucociliary clearance, and airway injury (6,7). Consequently, this leads to chronic productive cough, copious sputum production, recurrent respiratory exacerbations, and progressive lung damage. (6,7).

*P. aeruginosa* is the most common pathogen affecting bronchiectasis airways worldwide (8–12), and is associated with more rapid lung function decline, advanced lung lobe involvement, worse quality of life, and most concerningly, a 7-fold increased risk of hospitalisation and a 3-fold increased risk of death (8–12).With the global rise of AMR *P. aeruginosa* strains and occasional reports of pan-drug-resistant strains, treatment is becoming increasingly complicated for an already notorious and difficult-to-treat pathogen.

Multidrug (MDR) (i.e. resistance towards three or more antibiotic classes) and XDR (i.e. resistance to all except 1-2 classes) *P. aeruginosa* pose an increasing challenge in the treatment of chronic respiratory disease (13,14). MDR and XDR *P. aeruginosa* have been comprehensively studied in CF (14–19); however, there is a relative paucity of similar studies of bronchiectasis-derived isolates. Among the limited published studies, reported MDR rates vary considerably. For instance, similar MDR rates of 16.3%, 16%, and 11% were found in a 2026 Chinese study of 306 non-duplicate *P. aeruginosa* isolates, a small 2022 Spanish study of 43 *P. aeruginosa* isolates, and a retrospective 2015 UK study of 155 participants, respectively (20–22). A 2018 Chinese study of 147 *P. aeruginosa* isolates found a slightly higher MDR rate of 23.1%, which were associated with increased hospitalisation (23). In contrast, a 2025 Chinese study found that 84.2% of bronchiectasis-derived *P. aeruginosa* were carbapenem resistant, with many isolates (*n=*16/38, 42.1%) harbouring the high-risk, mobile, *bla*_OXA-50_ carbapenemase gene (24).

There is a relative dearth of AMR data available from bronchiectasis-derived *P. aeruginosa* in Australia. A recent study by Madden *et al.* found that Australian *P. aeruginosa* isolates from other chronic respiratory diseases can develop AMR in undetermined ways compared with comprehensively studied strains from Europe and the United States (25). Uncovering regional strain variations is essential for comprehensive epidemiologic understanding of bronchiectasis-derived *P. aeruginosa*. It is therefore a priority to better understand *P. aeruginosa* population genetics, AMR rates and associated genetic determinants, and persistence rates from bronchiectasis in understudied geographic regions such as Australia.

To address this knowledge gap, we performed antimicrobial susceptibility testing (AST), whole-genome sequencing (WGS), multilocus sequence typing (MLST), phylogenomics, and *in silico* AMR determinant identification (25,26) on 137 *P. aeruginosa* isolates retrieved from sputa of 59 Australians with bronchiectasis. We subsequently compared our strains alongside 4,575 previously published *P. aeruginosa* genomes from international bronchiectasis cohorts (27–34) to unmask global phylogeographic, within-host evolution, and AMR determinant patterns.

## METHODS

### Ethical approval

*P. aeruginosa* isolates (*n=*137) were obtained from sputa collected from Australians with computed tomography-confirmed bronchiectasis as part of retrospective (Bronchiectasis and Low-Dose Erythromycin Study [BLESS] [*n=* 42]) and prospective (BRONX [*n=*58], CERTAIN [*n=*13] or both [*n=*13]) studies. The BLESS Study was approved by the Mater Misericordiae Ltd Human Research Ethics Committee (HREC/MML/89908); the BRONX and CERTAIN Studies were approved by the Metro North Health Human Research Ethics Committee (HREC/2024/MNHB/107113 and HREC/2023/MNHB/90934, respectively). Site-specific approvals were subsequently obtained from the Research Governance Offices at each hospital site (Mater Hospital Brisbane, The Prince Charles Hospital, and Sunshine Coast University Hospital) prior to participant recruitment and *P. aeruginosa* isolation.

### Participant population

BLESS participants were previously recruited for a 12-month, double-blind, randomised, placebo-controlled trial that investigated low-dose erythromycin efficacy in adults with bronchiectasis presenting to Mater Hospital Brisbane (South Brisbane, QLD, Australia; Nov 2008-Oct 2010) (35,36), many of whom were re-recruited during the INFINITY Study (May 2017-Dec 2018) for follow-up. The CERTAIN Study was an interventional, non-randomised, unblinded clinical trial that investigated ceftolozane/tazobactam (C/T) efficacy for treating *P. aeruginosa* exacerbations in sputum-productive adults with bronchiectasis (including CF) who presented to Mater Hospital Brisbane, The Prince Charles Hospital (Chermside, QLD, Australia), or Sunshine Coast University Hospital (Birtinya, QLD, Australia) between Jan 2024 and Dec 2025 (Australian New Zealand Clinical Trials Registry [ACTRN12623000976684]). The BRONX Study was an exploratory research study that recruited sputum-productive adults with bronchiectasis presenting at the Sunshine Coast University Hospital between Jan 2024 and Dec 2025. Due to the varying structure and protocol of each trial, some participants provided longitudinal sputa, whereas others provided a singular timepoint. Sputum timepoints, antibiotic regimen, demographic data, and study of enrolment for each participant were collated (Table S1). All patients performed spirometry at baseline, with BLESS participants re-performing spirometry during the follow-up INFINITY Study.

### P. aeruginosa isolation

BLESS and INFINITY bronchiectasis sputa, which had been stored for several years at -80°C without cryoprotectant, were plated onto Luria-Bertani (LB) agar (Becton, Dickinson and company, MD, USA) at 37°C with 5% CO_2_ for 24-48h, which resulted in good (43/68 sputa; 63.2%) *P. aeruginosa* viability. Prospectively collected BRONX and CERTAIN Study sputa were plated aerobically onto MacConkey agar (Becton Dickinson Pty Ltd, Macquarie Park, NSW, Australia) within 48h of sputum receipt, with any non-lactose-fermenting colonies subcultured for speciation. All plates were incubated at 37°C for 24± 2h.

### *P. aeruginosa* species confirmation

DNA was extracted from pure cultures using 5% Chelex-100 (Bio-Rad Laboratories Pty Ltd, South Granville, NSW, Australia), followed by a 10 min incubation at 95°C to lyse cells. 1:10 diluted supernatant was screened in duplicate for *P. aeruginosa* presence and panbacterial load using an *ecfX* (*PA1300*)-16S rRNA duplex real-time PCR assay targeting *P. aeruginosa* (37) and all bacteria (38), respectively, using previously described parameters (39). At least one positive control (*P. aeruginosa* strain SCHI0005.S.8; NCBI accession: SRR15793216) and one reagent-only no-template control (NTC) were included on each PCR run.

### Antibiotic susceptibility testing

Susceptibility profiles were determined for eleven antipseudomonal antibiotics. Ten antibiotics were tested by disc diffusion (Edwards Group, Murrarie, QLD, Australia) (40) using cation-adjusted Mueller-Hinton agar (Becton Dickinson): amikacin (AMK; 30 mg), aztreonam (ATM; 30 mg), cefepime (FEP; 30 mg), ceftazidime (CAZ; 30 mg), ceftolozane/tazobactam (C/T; 30/10 mg), ciprofloxacin (CIP; 5 mg), imipenem (IPM; 10 mg), meropenem (MEM; 10 mg), piperacillin/tazobactam (TZP; 100/10 mg) and tobramycin (TOB; 10 mg). As chemical properties limit its diffusion through agar (41), colistin (CST) susceptibility was determined by broth microdilution. Isolates were classified as susceptible, susceptible (increased exposure), or resistant using the EUCAST v 16.1 2026 breakpoints (42) for AMK, ATM, FEP, C/T, CIP, IPM or MEM, and using the CLSI M100 35^th^ edition (43) for CAZ and TZP. The AMR-sensitive control strain PAO1 (LMG 12228) was used as a control strain for all testing to confirm antibiotic integrity.

Definitions of AMR, MDR, XDR, and pan-drug resistance were based on the European Centre for Disease Prevention and Control and US Centers for Disease Control and Prevention guidance (13). Briefly, isolates were classified as AMR when resistant towards 1-2 antimicrobial categories, MDR when resistant to at least one agent in three or more antimicrobial categories, XDR when resistant to at least one agent in all except one or two antimicrobial categories, and pan-drug resistant when resistant towards all agents in all categories (13).

### Whole-genome sequencing

Genomic DNA was extracted from purified isolates by transferring a ∼10 μL loopful of agar-cultured cells into 200 μL lysis buffer (20 mM Tris-HCl pH 8.0, 2 mM sodium EDTA, and 1.2% Triton X-100 [Sigma-Aldrich]), followed by bead beating in DNase-free 2 mL O-ring tubes (SSIBio, Lod, CA, USA) containing ∼100 μL of 0.1 and 0.5 mm zirconia beads (Daintree Scientific, St Helens, TAS, Australia) using a Precellys Evolution Touch Homogeniser (Bertin Technologies, Montigny-le-Bretonneux, France) at 2 x 7500 rpm for 60 sec (with a 30 sec pause between cycles). DNA was purified using the ‘Gram-positive Bacteria’ protocol of the DNeasy Blood and Tissue kit (QIAGEN, Clayton, VIC, Australia), commencing at the Buffer AL and Proteinase K addition step. Extracted DNA was quantified using a Qubit 4 fluorometer (Thermo Fisher Scientific, Scoresby, VIC, Australia). WGS was performed using Illumina NovaSeq X to generate 150bp paired-end reads at ∼100x depth (AZENTA, Suzhou, China and Macrogen, Geumcheon-gu, Seoul, Korea).

### Bioinformatic analyses

Following genome quality filtering and assessment using Trimmomatic v0.39 (44) and FastQC v12.1 (45), species identification was conducted using Kraken 2 v2.17.1 (46). AMR-conferring determinants were identified using ARDaP v2.3.2 (25,26). For variant analysis, SPANDx v4.2 (47) was used to first quality-filter reads and then detect single-nucleotide polymorphisms (SNPs) and insertions/deletions (indels). SPANDx-generated SNP matrices were used to construct phylogenetic trees using FastTree v2.2.0 (46); trees were subsequently visualised using iTOL v7.4.2 (48). Genome assembly was undertaken using Microbial Genome Assembler Pipeline v3.5 (49), with PAO1 (GenBank ref: AE004091.2) included as the scaffolding reference. Assembled contigs were then aligned against PAO1 and annotated with Prokka v1.15.6 (50). *In silico* multilocus sequence typing (MLST) was performed on assembled genomes using the *P. aeruginosa* PubMLST database (available at https://pubmlst.org/organisms/pseudomonas-aeruginosa), with alleles and STs assigned via the embedded BIGSdb software (51). Recombination was examined using Gubbins v3.4.1 (52). All assembled genomes were uploaded to GenBank (BioProject ID PRJNA1325058) and *P. aeruginosa* PubMLST databases, and Illumina reads were uploaded to the NCBI SRA database (BioProject IDs PRJNA1330417 and PRJNA1325058).

### Publicly available *P. aeruginosa* genomes from international bronchiectasis datasets

Data were downloaded for 4,575 *P. aeruginosa* isolates from eight published (27–34) and two unpublished (BioProject: PRJNA1419033 and PRJNA1376148) studies. International isolates were from the UK (*n=*185, 4.0%), Germany (*n=*117, 2.5%), Spain (*n=*46, 1.0%), China (*n=*31, 0.6%), and USA (*n=*5, 0.1%), with most of unknown origin (*n=*4201, 91.8%). These data were integrated with our 137 *P. aeruginosa* bronchiectasis isolates and bioinformatically analysed as described above.

### Statistical analysis

Significant associations between isolate genotypes, phenotypes, and participant clinical features were assessed using Microsoft Excel v 16.104. A chi-square test of independence (CHISQ.TEST function) was used to assess the association between categorical variables (e.g. *P. aeruginosa* AMR classification versus participant lung function severity). Differences in continuous variables (age, body mass index [BMI], bronchiectasis severity index [BSI], and forced expiratory volume in one second [FEV₁]) between males and females were assessed using unpaired *t*-tests. Statistical significance was defined as *p*<0.05. The SNP accumulation rate (i.e. SNPs-per-year rate) was calculated using R v4.6.0 (2026-04-24 ucrt) via the lm() function, with data plotted and visualised in RStudio (v2026.06.0 Build 242).

## RESULTS

### Demographic data

Our cohort represents the typical bronchiectasis population, with elderly, female, non-smokers being most common, reflecting findings from the Australian Bronchiectasis Registry (8,53) (Table 1). Average participant age was 67.8 ± 14.0 years with most participants being female (*n=*44, 74.6%). Average BMI was borderline normal-overweight at 25.7kg/m^2^ ± 8.0 (15 – 65kg/m^2^), and most were never smokers (*n=*44, 74.6%). As per ATS/ERS 2005 guidelines (54), average lung function was classified as moderate, with an average FEV_1_ of 60.3 ± 22.5% predicted. Of those with Bronchiectasis Severity Scores (BSI) available, most (*n=*35/53, 66.0%) had severe BSI (i.e. score >9/20), with 24.5% (*n=* 13/53) having a BSI score ≥15/20. Bronchiectasis aetiology was predominantly post-infective (36.2%), followed by idiopathic (12.1%), mercury associated (8.6%), and other sub-categories (43%) (Table 1). There were no statistically significant differences between male and female demographic variables based on age, smoking status, BMI, BSI score, or FEV_1_ (*p>*0.05).

**Table 1.**
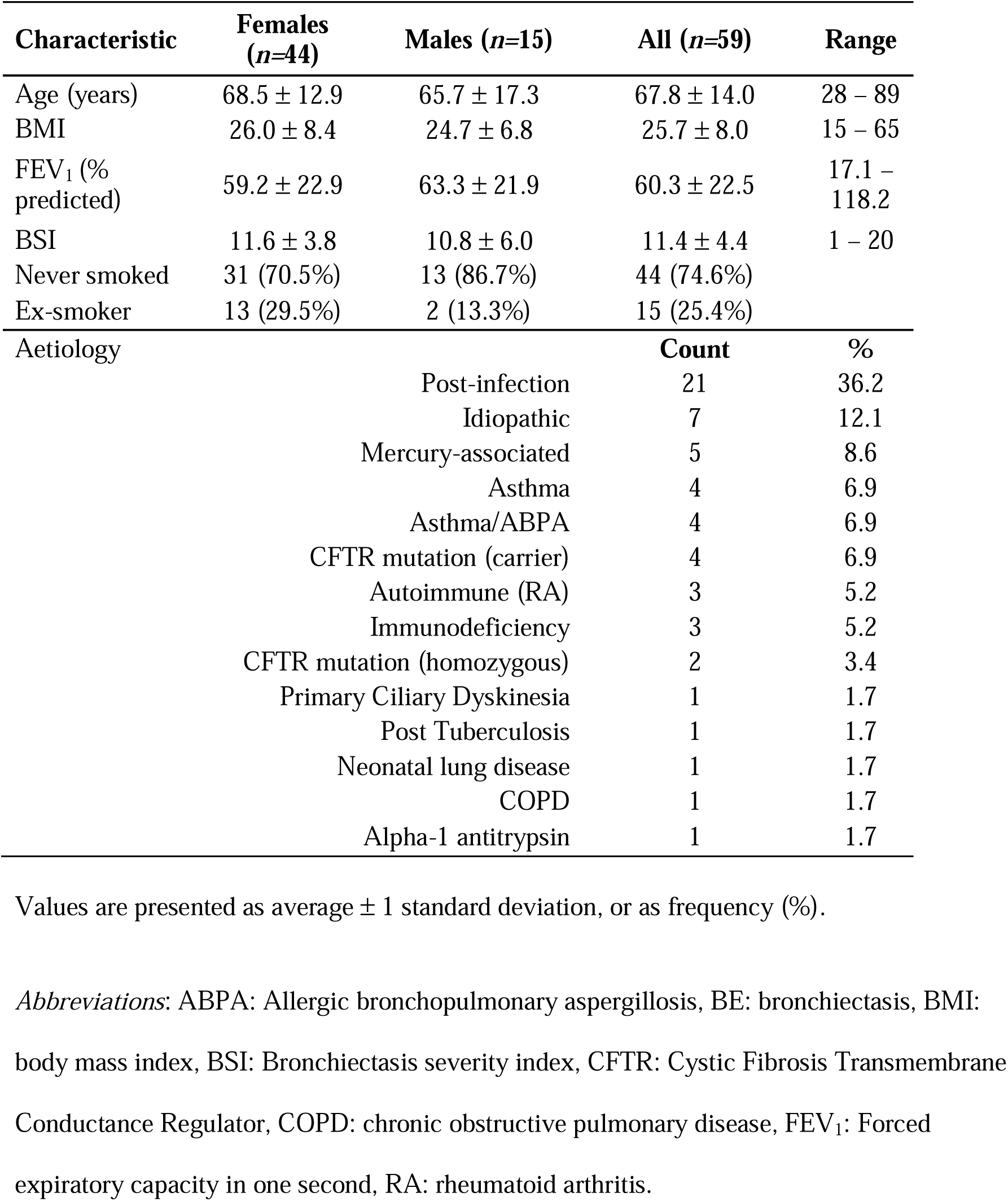
Demographic details of bronchiectasis participants.

| **Characteristic** | **Females (*n*=44)** | **Males (*n*=15)** | **All (*n*=59)** | **Range** |
|---|---|---|---|---|
| Age (years) | 68.5 ± 12.9 | 65.7 ± 17.3 | 67.8 ± 14.0 | 28 – 89 |
| BMI | 26.0 ± 8.4 | 24.7 ± 6.8 | 25.7 ± 8.0 | 15 – 65 |
| FEV<sub>1</sub> (% predicted) | 59.2 ± 22.9 | 63.3 ± 21.9 | 60.3 ± 22.5 | 17.1 – 118.2 |
| BSI | 11.6 ± 3.8 | 10.8 ± 6.0 | 11.4 ± 4.4 | 1 – 20 |
| Never smoked | 31 (70.5%) | 13 (86.7%) | 44 (74.6%) |  |
| Ex-smoker | 13 (29.5%) | 2 (13.3%) | 15 (25.4%) |  |
| Aetiology |  |  | **Count** | **%** |
| Post-infection |  |  | 21 | 36.2 |
| Idiopathic |  |  | 7 | 12.1 |
| Mercury-associated |  |  | 5 | 8.6 |
| Asthma |  |  | 4 | 6.9 |
| Asthma/ABPA |  |  | 4 | 6.9 |
| CFTR mutation (carrier) |  |  | 4 | 6.9 |
| Autoimmune (RA) |  |  | 3 | 5.2 |
| Immunodeficiency |  |  | 3 | 5.2 |
| CFTR mutation (homozygous) |  |  | 2 | 3.4 |
| Primary Ciliary Dyskinesia |  |  | 1 | 1.7 |
| Post Tuberculosis |  |  | 1 | 1.7 |
| Neonatal lung disease |  |  | 1 | 1.7 |
| COPD |  |  | 1 | 1.7 |
| Alpha-1 antitrypsin |  |  | 1 | 1.7 |

Values are presented as average ± 1 standard deviation, or as frequency (%).
*Abbreviations*: ABPA: Allergic bronchopulmonary aspergillosis, BE: bronchiectasis, BMI: body mass index, BSI: Bronchiectasis severity index, CFTR: Cystic Fibrosis Transmembrane Conductance Regulator, COPD: chronic obstructive pulmonary disease, FEV<sub>1</sub>: Forced expiratory capacity in one second, RA: rheumatoid arthritis.

### MLST

Fifty-seven sequence types (ST) were identified in our dataset (Table S3), including seven novel STs (ST5013, ST5014, ST5015, ST6238, ST6239, ST6243, ST6966). The high-risk international clone ST274 (*n=*9, 6.6%) and ST2128 (*n=*8, 5.8%) were most common, followed by ST253 (*n=*7, 5.1%), ST655 (*n=*7, 5.1%), ST170 (*n=*5, 3.6%), and the common global clone ST155 (*n=*5, 3.6%). Although less frequent, other notable STs included the high-risk international clones ST395 (*n=*1, 0.7%), ST244 (*n=*1, 0.7%), ST277 (*n=*1, 0.7%), ST298 (*n=*1, 0.7%), and common global clones ST782 (*n=*2, 1.4%), ST27 (*n=*1, 0.7%), and ST260 (*n=*1, 0.7%) (Figure 1).

**Figure 1.**
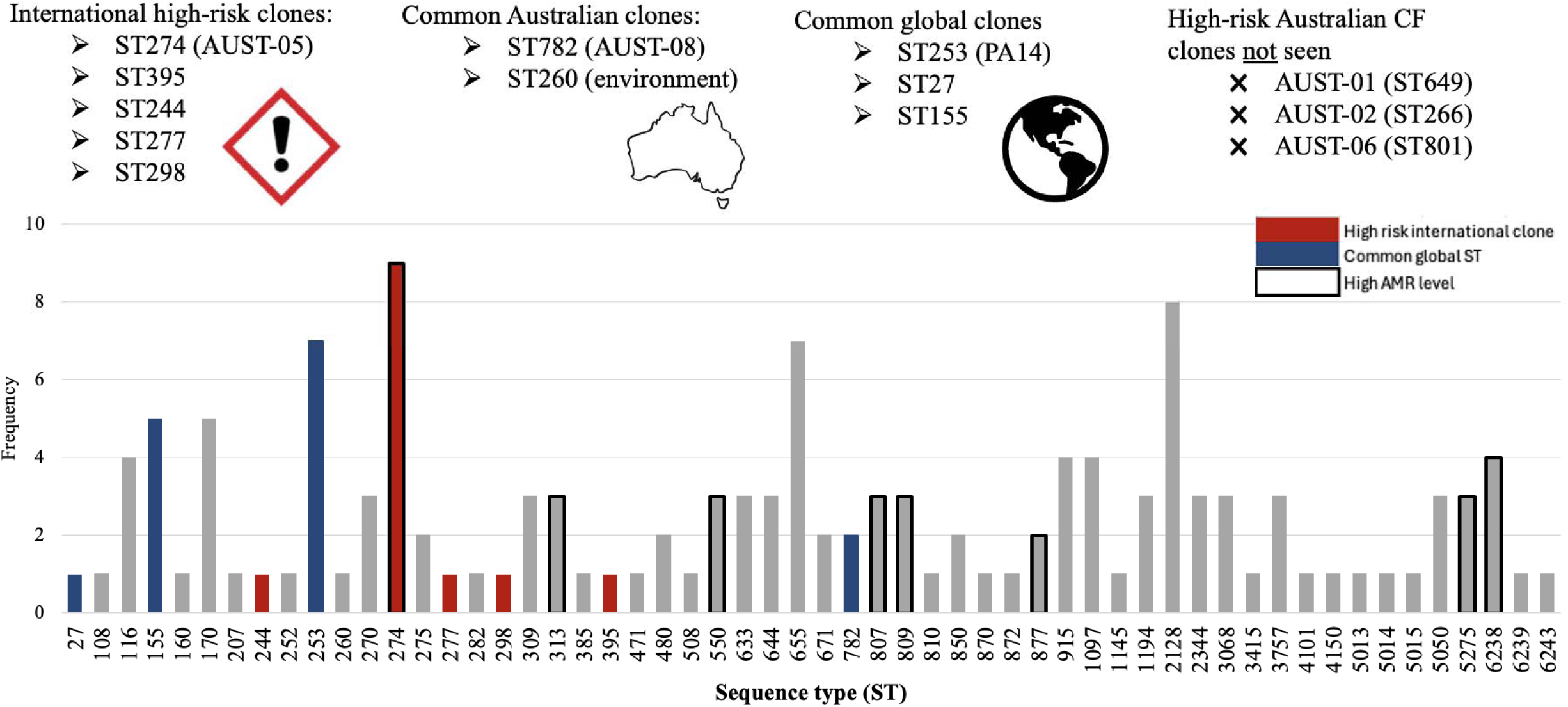
Multilocus sequence type (ST) prevalence and associated antimicrobial resistance (AMR) rates in our bronchiectasis cohort.

### Antimicrobial susceptibility testing

AMR rates were high, with 75.9% (*n=*104/137) isolates being resistant to at least one clinically relevant antibiotic. Specifically, 46.7% (*n=*64/137) were AMR, 19.7% (*n=*27/137) were MDR, and 9.5% (*n=*13/137) were XDR (i.e. resistant to all except one or two categories (13,42)), with the remaining isolates (24.1%; *n*=33/137) sensitive to all tested antimicrobials (Table S3; Figure 2). No pan-drug resistant isolates were observed. CIP resistance was most common (*n=*88, 64.2%), followed by FEP (*n=*61, 44.5%), C/T (*n=*40, 29.2%), ATM (*n=*29, 21.2%), CAZ (*n=*27, 19.7%), IPM (*n=*26, 19.0%), MEM (*n=*21, 15.3%), AMK (*n=*16, 11.7%), TOB (*n=*14, 10.2%), TZP (*n=*12, 8.8%), and CST (*n=*1, 0.7%) resistance (Figure 2). AMR/MDR/XDR classification was not associated with lung function severity (*p=*0.783); however, AMR evolved over time for several participants (Table S3). Phenotypically, 56.2% (*n=*77/137) isolates displayed a mucoid phenotype (Table S3).

**Figure 2.**
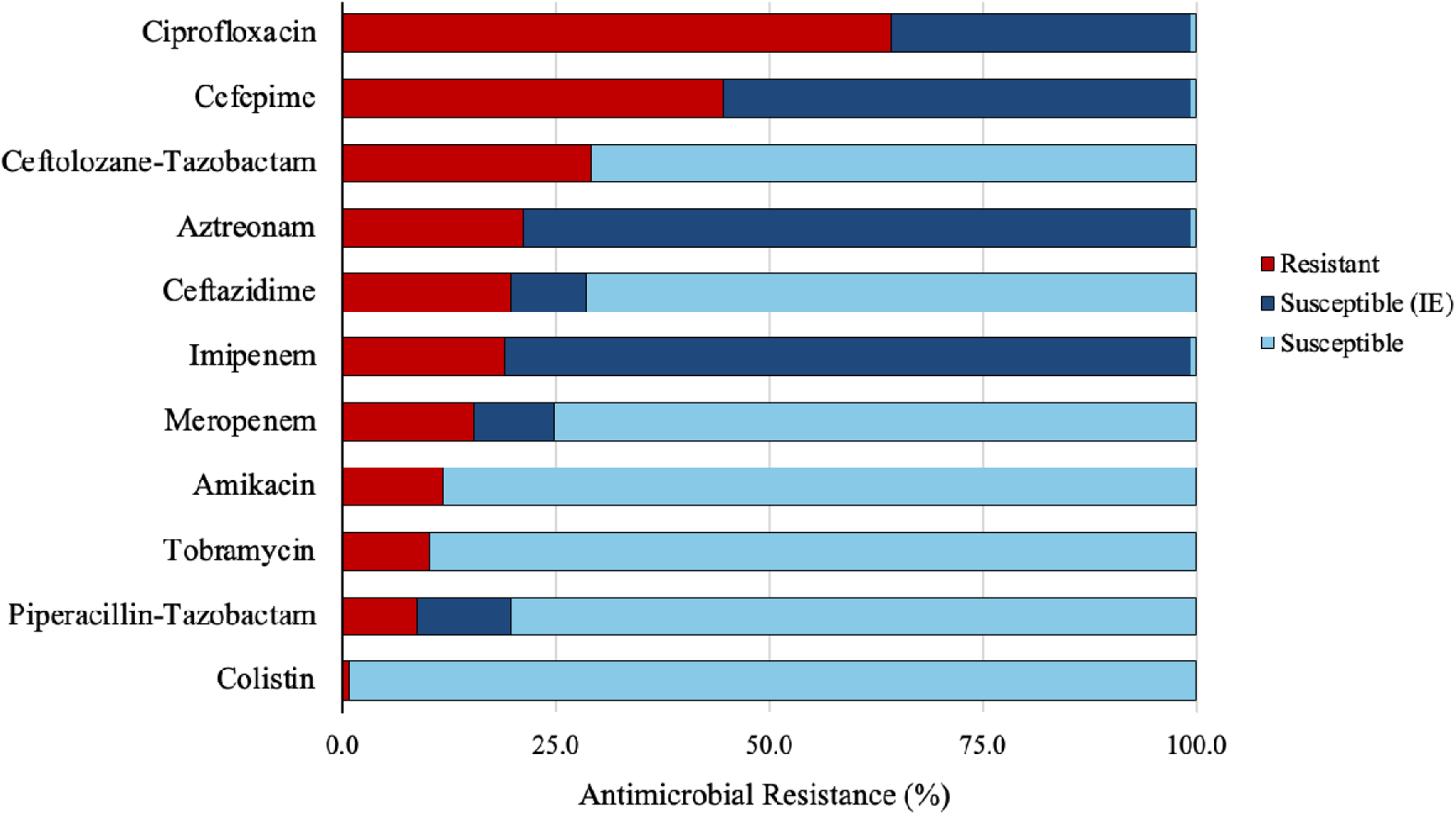
Antibiotic susceptibility rates of 137 Australian *Pseudomonas aeruginosa* isolates derived from bronchiectasis airways against 11 antipseudomonal antibiotics. NB. Under EUCAST 2026 guidelines, wild-type *P. aeruginosa* isolates are classified as ‘susceptible, increased exposure (IE)’ for ciprofloxacin, cefepime, aztreonam and imipenem. The standard ‘susceptible’ category is effectively removed for these drugs by setting an arbitrary breakpoint of >50mm (42). Therefore, the high proportion of ‘susceptible, IE’ isolates in our collection represents baseline, wild-type *P. aeruginosa* strains lacking acquired resistance determinants. Clinically, this classification mandates that *P. aeruginosa* eradication using these drugs requires optimised pharmacokinetic exposures, such as increased dosing, prolonged infusions, or a more targeted delivery route.

### *In silico* AMR profiling

Several known and novel AMR determinants were identified *in silico* in our cohort (Table 2). AMR determinants correlated well with phenotypic resistance, with genomic drivers of AMR identified in most resistant isolates.

**Table 2.** Key AMR mutations identified in antimicrobial-resistant *Pseudomonas aeruginosa* isolates retrieved from bronchiectasis airways.

| Antibiotic class and agent | Gene | Key Mutation/s identified |
|---|---|---|
| ***Fluoroquinolones*** |  |  |
| • Ciprofloxacin | *gyrA* | Asp72Gly, Thr83Ala/Ile/Val, Asp87Asn/Tyr |
|  | *gyrB* | Ser466Phe, Glu468Asp |
|  | *parC* | Ser87Leu |
|  | *mexT* | Loss (including Gln80fs) |
|  | *nfxB* | Ter188Cysext*? |
| ***β lactams*** |  |  |
| • Cephalosporins +/- inhibitor (ceftazidime, cefepime, ceftolozane/tazobactam), Carbapenems (meropenem, imipenem), Penicillin + inhibitor (piperacillin/tazobactam) | *ampC* | Val239Ala |
|  | *ampD* | Val10Gly, Gly12fs |
|  | *ampR* | Arg86Cys, Asp135Asn |
|  | *argJ* | Loss (including Gly88fs, Ala91fs, Ala90_Ala91insProPro) |
|  | *dacB* | Loss (including Ala217fs, Leu259fs) |
|  | *ftsI* | Gly63Asp |
|  | *mrcB* | Loss |
|  | *oprD* | Loss (including Phe36fs, Trp65*, Gln79fs, Gln166fs, Phe308fs, Arg342fs, Trp417*, Ala420fs) |
|  | *PA0667* | Loss (including Arg334fs, Gly337fs, Tyr338fs) |
|  | *PA4677* | Asp138Val |
|  | *pepP* | Loss |
| ***Aminoglycosides*** |  |  |
| • Amikacin, tobramycin | *fusA1* | Gly545Asp |
| ***Polymyxin*** |  |  |
| • Colistin | *pmrB* | Val136Met |
|  | *phoQ* | Gln263His |

β-lactam resistance determinants were heterogenous in nature, with numerous mutation combinations responsible for conferring resistance (Table S2). Mutations typically occurred in genes controlling AmpC regulation (*ampR, dacB*) and structure (*ampC*), penicillin-binding protein production (PBPs; *ftsI, dacB*), outer-membrane porin structure (*oprD*), or efflux pump regulation (*mexR, mexT, nalD*). Missense mutations in the transcriptional regulatory gene, *ampR*, were seen in 7.3% (*n=*10/137) of isolates (Table S4), most of which were classified as MDR or XDR (Table S3). We also identified four nonsense *oprD* variants affecting residues 1-82 (Table S3). Nine (6.5%) isolates were resistant to all four β-lactam classes tested (carbapenems, cephalosporins, penicillins, monobactams), all of which had multiple mutations across several β-lactam-associated genes. For example, SCHI0181.S.68, which was resistant to all β-lactams, including carbapenems and cephalosporins (Table 2), encoded deleterious mutations in *ftsI* (likely reducing β-lactam affinity to PBP3), *nalD* (likely causing efflux pump upregulation), *oprD* (likely reducing cell permeability), and *ampR* (likely resulting in *ampC* upregulation) (Table S3).

To evaluate the efficacy of β-lactam/β-lactamase inhibitor combinations, we compared TZP resistance rates against C/T, a newer antimicrobial specifically engineered to evade AmpC hydrolysis and efflux (55,56). Unexpectedly, C/T resistance (29.2%, *n=*40/137) was over three-fold higher than TZP resistance (8.8%, *n=*12/137), a trend that remained even when accounting for multiple isolates from individual participants (37.3%, *n=*22/59 participants vs. 10.1%, *n=*6/59 participants). Interestingly, TZP resistance was significantly associated with XDR status (*p=* <0.001). In contrast, C/T resistance was not significantly associated with active antibiotic administration, resistance to other compounds, or AMR/MDR/XDR status, suggesting the presence of intrinsic resistance with distinct AMR mechanisms. Although we observed specific *ampR* and/or *ampC* variants in some C/T resistant isolates, the AMR driver mutation remained unknown for most isolates.

We unexpectedly identified a CST-resistant *P. aeruginosa* isolate, SCHI0273.S.1, from P43, who was receiving long-term nebulised CST at the time of sputum collection. To facilitate comparative genomic analysis, we attempted to retrieve CST sensitive strains from this participant’s sputum; however, all >70 subcultured clones were CST resistant, indicating a likely fixed population. Closer inspection of SNP and indel variants identified two missense mutations affecting PmrB (Val136Met) and PhoQ (Gln263His) in SCHI0273.S.1 (Table S3). These mutations were not identified in strains obtained from any other participant.

### Within-host evolution

Among our longitudinal cases, phylogenomic reconstruction revealed a strong signal of within-host evolution in participants who were infected for at least 6 months (*n=*13/13 [100%]). Within-host genetic distances ranged from 8 to 1646 accumulated SNP differences (Table S3), with an average of 1.16 substitutions per million base pairs/year, excluding two outliers (Figure 4).

In most cases, AMR rates did not worsen over time, consistent with the initial isolates often already possessing high AMR levels, and in some cases (P06, P39, P45), isolates became more susceptible over time. Of the participants who developed worsening AMR, we identified several chromosomal AMR determinants associated with acquired phenotypic resistance. The most common AMR determinants in our longitudinal cohort were GyrA Thr83Ile/Val and Asp87Asn/Tyr (CIP resistance), AmpR Arg86Cys and Asp135Asn (cephalosporin resistance), and nonsense OprD (carbapenem resistance), occurring in two or more participants each.

In P09, >1,000 SNPs accumulated in the latter isolates over a nearly nine-year period, including mutations affecting MexT (Gln80fs) and GyrA (Thr83Ile) that drove MDR development (Table S3). Additionally, all 2018 isolates retrieved from P09 underwent genome reduction, losing a total of 56.1kb of the core genome (a 52.3kb deletion affecting *PA0672*-*PA0714* [coordinates 733074-785336 in PAO1], and a 3.8kb deletion affecting *PA4307*-*PA4309* [coordinates 4832685-4836526 in PAO1]) compared to the initial 2009 isolate. Impacted loci encode pathways involved nutrient scavenging, stress-responsive secretion systems, virulence, and cell envelope modifications, including the Vre virulence factor transcription system (*PA0673*–*PA0676*), Hxc Type II secretion system (*PA0677*-*PA0687*), chemotaxis (*PA4307-PA4309*), iron acquisition (*hemO*, *exbB2*, *exhD2*), and lipopolysaccharide production (*migA*).

Examination of within-host evolution signatures across all participants unveiled the most common loss-of-function mutations in genes encoding motility and attachment (structural flagellar and Type IV pili components such as *pilA*, *pilB*, *pilW*, *fliC*; 640 total mutations), lipopolysaccharide modification (e.g. *wzy*, *wbpL*, *wbpM* and *rmlB* O-antigen synthesis and *pagL* lipid A modification loci; 345 total mutations), virulence and quorum sensing (e.g. *lasR* and *PA5438* master quorum-sensing regulators, *exoS* and *exoY* Type III Secretion System effectors or components; 145 total mutations), biofilm, exopolysaccharide, and mucoidy production (*mucA* or *mucD*-associated alginate overproduction; 106 mutations), and energy-intensive iron and haem acquisition machinery (e.g. *pchF*, *pchE*, *hemO*; 93 total mutations) (Table S3, Table S5). Furthermore, twenty-eight strains encoded the PA5438 (GavR) Ser275_Arg277del variant, which has been associated with chronic *P. aeruginosa* infection establishment (32).

Multilineage infections (co-infection with two or more genetically distinct isolates) were detected in 12.5% (*n=*4/32) of participants (Figure 3; Table S3). In addition to multilineage infections, comparative genomic analysis identified that two longitudinal infection cases, P05 and P09, harboured isolates that likely changed STs within-host due to single missense variants in MutL (Arg264Cys) and AroE (Lys218Arg), respectively, becoming single-locus variants (Table S3).

**Figure 3.**
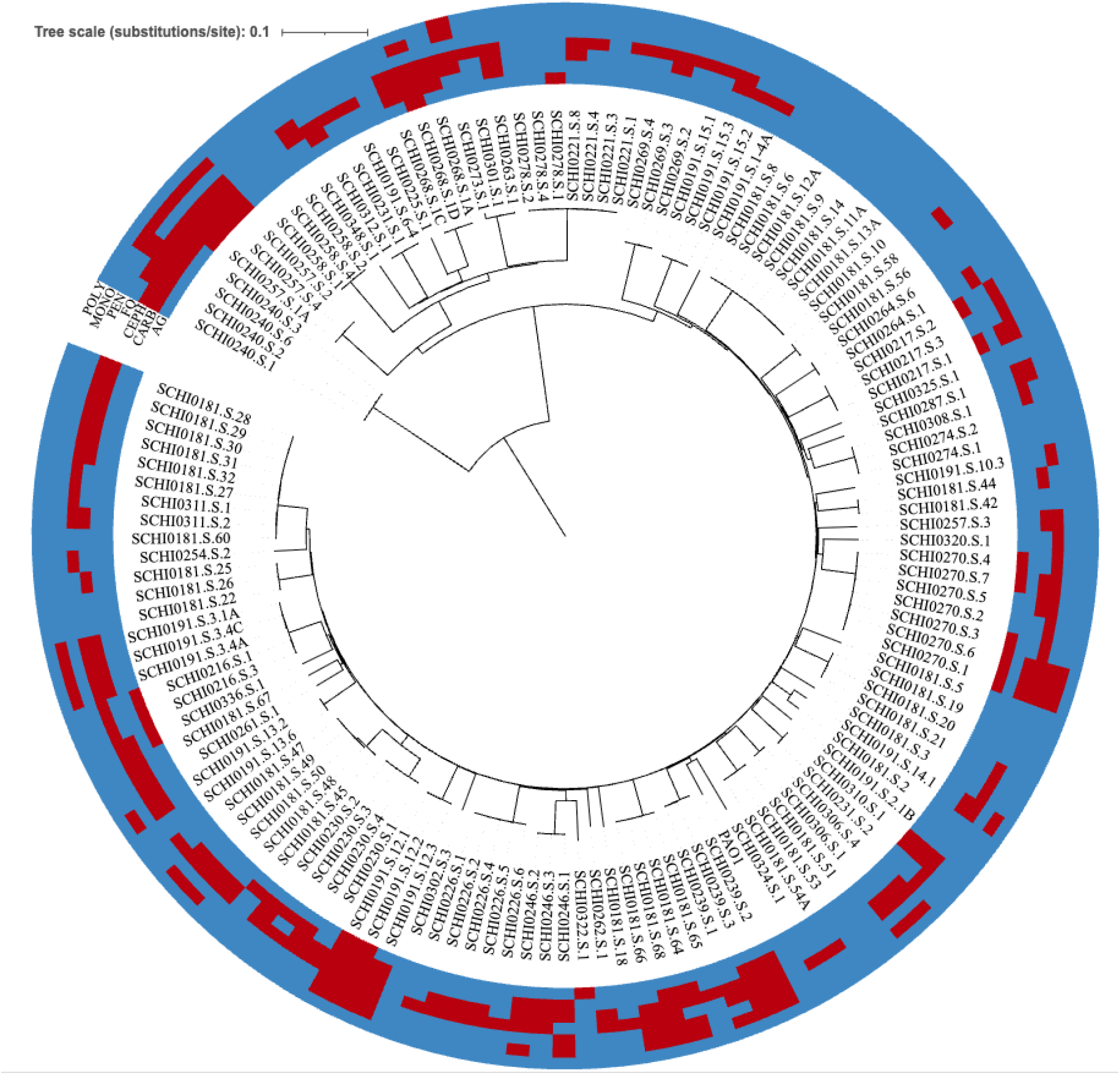
Midpoint-rooted maximum parsimony phylogenomic analysis of 137 Australian bronchiectasis lung-derived *P. aeruginosa* isolates. Single-nucleotide polymorphisms were identified using SPANDx v4.2 (47) and the phylogeny reconstructed using FastTree v2.2.0 (46). Antibiotic class susceptibility profile is shown in the outer rings (red: resistant, blue: susceptible [including susceptible, increased exposure). PAO1 was used as the read mapping reference. Phylogenetic analysis visualised using iTOL v 7.4.2 (48). Abbreviations: AG: aminoglycosides; CARB: carbapenems; CEPH: cephalosporins; FQ: fluoroquinolones; PEN: penicillins; MONO: monobactam; POLY: polymyxin; substitutions/site: nucleotide substitutions per site.

### Hypermutation

A common phenomenon in CF-derived *P. aeruginosa* is hypermutator emergence, especially in chronic infections (36-60% incidence) (57–59). Hypermutators lack DNA repair mechanisms, typically due to high-consequence defects in *mutS* and *mutL* DNA mismatch repair systems, resulting in up to 1000-fold higher spontaneous mutation rate compared to wild-type strains (57–59). Hypermutation has also been reported in the *P. aeruginosa*-positive bronchiectasis population, albeit less frequently (∼9%) than in CF (27). We similarly found that 13.1% (*n=*18) of our isolates encoded high-consequence *mutS* (*n=*10; 7.3%) or *mutL* (*n=*8; 5.8%) mutations (P04: MutL Glu205*, P30: MutS Leu771fs, P31: MutS Cys295fs, P40: MutL Ile475_Arg479del, P41: MutL Ala347fs, P46: MutL Thr402del, P47: MutS Gly422fs, P49: MutS Thr115fs, P56: MutS Trp851*. Table S3). When assessing hypermutated longitudinal isolates spanning at least six months, we observed an average rate of 24.5 substitutions per million base pairs/year compared with the average of 1.16 substitutions per million base pairs per year in non-hypermutated strains (i.e. a 21x higher mutation rate; Table S3). In four participants encoding high-consequence MutS (Gly422fs, Trp851*) or MutL (Glu205*, Thr402del) variants, only a single isolate was retrieved, hampering confirmation of their hypermutation status.

We also observed elevated mutation rates in strains lacking high-consequence MutS and MutL variants. P25, who presented with a persistent infection over the sampled 5-month timeframe, harboured two latter strains, SCHI0221.S.4 and SCHI0221.S.8, that had unusually accumulated >2,000 mutations, yet no hypermutation variants were identified (Table S3; Figure 4). Gubbins analysis confirmed a single recombination event in these latter strains, involving ∼55kb of the *P. aeruginosa* chromosome (SCHI0221.S.8 coordinates 5814054 to 5815135 [42 SNPs], 6100492 to 6121243 [658 SNPs] and 6130266 to 6163717 [1306 SNPs]), likely donated from an unsampled isolate co-infecting this participant. Similarly, P09 exhibited very high mutational burden over an 8.7-year timeframe (Figure 4); although no recombination or high-consequence hypermutation variants were identified (Table S3), a missense MutS Asp552Asn variant may explain the elevated SNP rate in these isolates.

**Figure 4.**
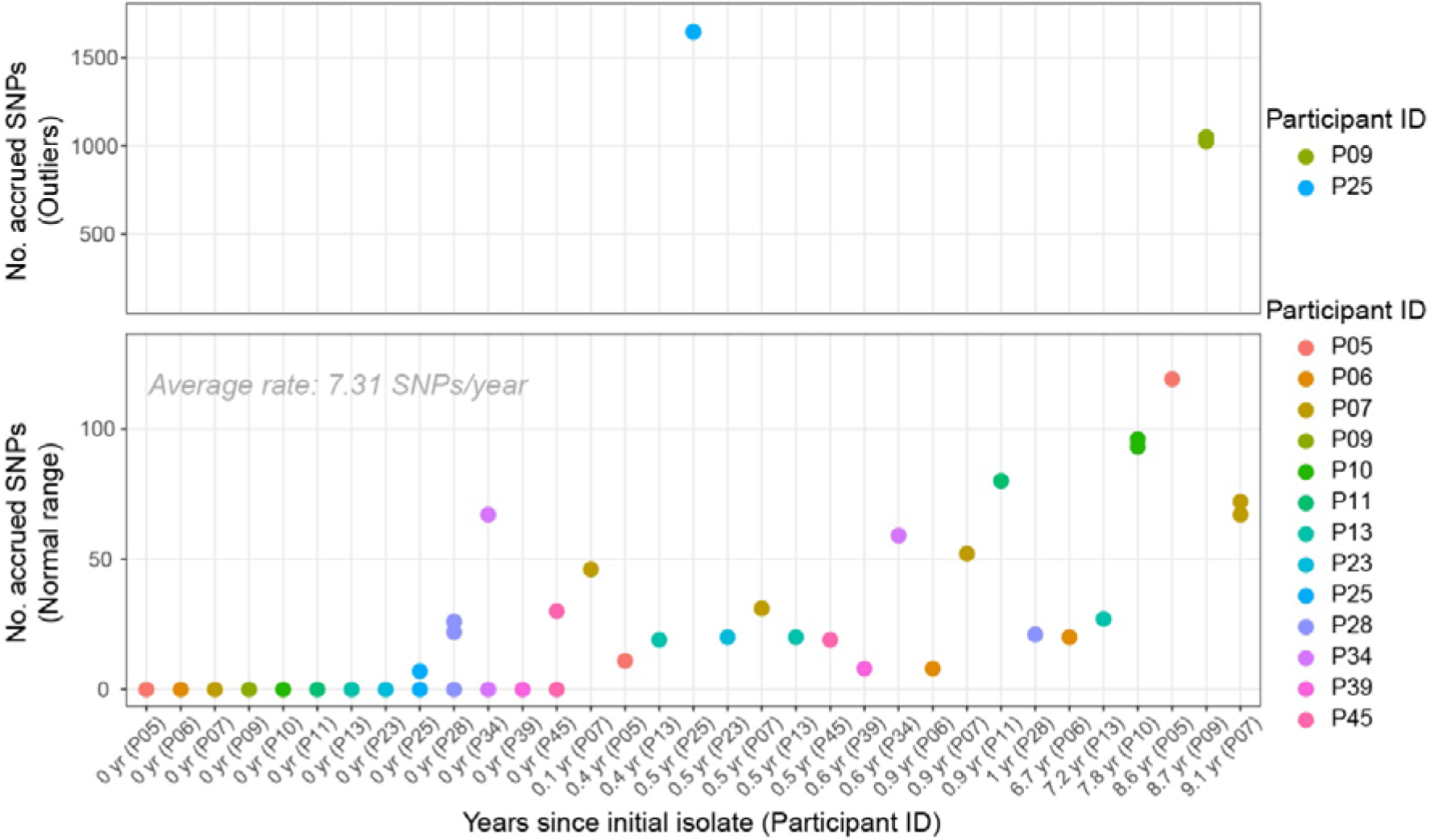
Single-nucleotide polymorphism (SNP) accumulation in longitudinal *Pseudomonas aeruginosa* obtained from bronchiectasis participants harbouring a persistent infection (≥6 months) with the same strain according to multilocus sequence typing. Among the 13 participants, two (P09 and P25) exhibited unusually high SNP differences between early and latter isolates. In P25, this phenomenon was attributed to a ∼55Kb recombination event; in P09, the cause was likely a *mutS* mutation (Asp552Asn) in the latter strains. Excluding outliers, our analysis identified that chronic *P. aeruginosa* infections in bronchiectasis airways accumulate an average of 7.31 SNPs/year, equating to 1.16 substitutions per million base pairs/year.

### Global isolate analysis

Bioinformatic analysis of all 4,575 publicly available bronchiectasis-derived *P. aeruginosa* genomes identified numerous known AMR determinants, with mutations in *mexB* (36.7%, *n=*1680/4575), *gyrA* (31.1%, *n=*1422/4575) and *mexA* (11.0%, *n=*504/4575) being most frequent (Table S4). This global AMR determinant prevalence was mostly consistent with our Australian cohort (Table S4). No significant differences were identified for the well-characterised GyrA AMR determinants Asp87His/Asn/Tyr (11.0% [our cohort] vs. 7.3% [global dataset]; *p*= 0.179), and Thr83Ile (24.1% vs. 21.5%; *p*= 0.503) Interestingly, our cohort had significantly higher prevalences of AmpC Val239Ala (4.4% vs 1.0%, *p <*0.001), AmpR Asp135Asn (5.1% vs 2.3%, *p=*0.029) and ArgJ loss (13.9% vs. 0.7%; *p*= <0.001), all of which encode resistance towards penicillins and/or cephalosporins. Five MGEs were identified in the global isolate genomes, all rare: two aminoglycoside modifying enzyme genes *aac(6’)-Ib-cr* (*n*=7/4575, 0.15%) and *ant(2’’)-Ia* (*n=*12/4575, 0.26%), two β-lactamase genes *bla*_oxa-10_ (*n=*11/4575, 0.24%) and *bla*_per-1_ (*n=*17/4575, 0.37%) and a plasmid-mediated quinolone resistance gene *qnrb19* (*n=*3/4575, 0.07%). No MGEs were identified in our local dataset.

Phylogenomic inclusion of our Australian isolates within the global dataset showed no distinct clustering based on geography, with Australian isolates interspersed among the global collection (Figure 5). However, country of origin data was unpublished for most global isolates (*n=*4201, 91.8%) (28), confounding our analysis. The isolates with available geographic origin data were from the UK (*n=*185, 4.0%), Germany (*n=*117, 2.5%), Spain (*n=*46, 1.0%), China (*n=*31, 0.6%), and USA (*n=*5, 0.1%). Using the available metadata, there was no strong evidence of phylogeographic clustering, suggesting a highly panmictic population (Figure 5).

**Figure 5.**
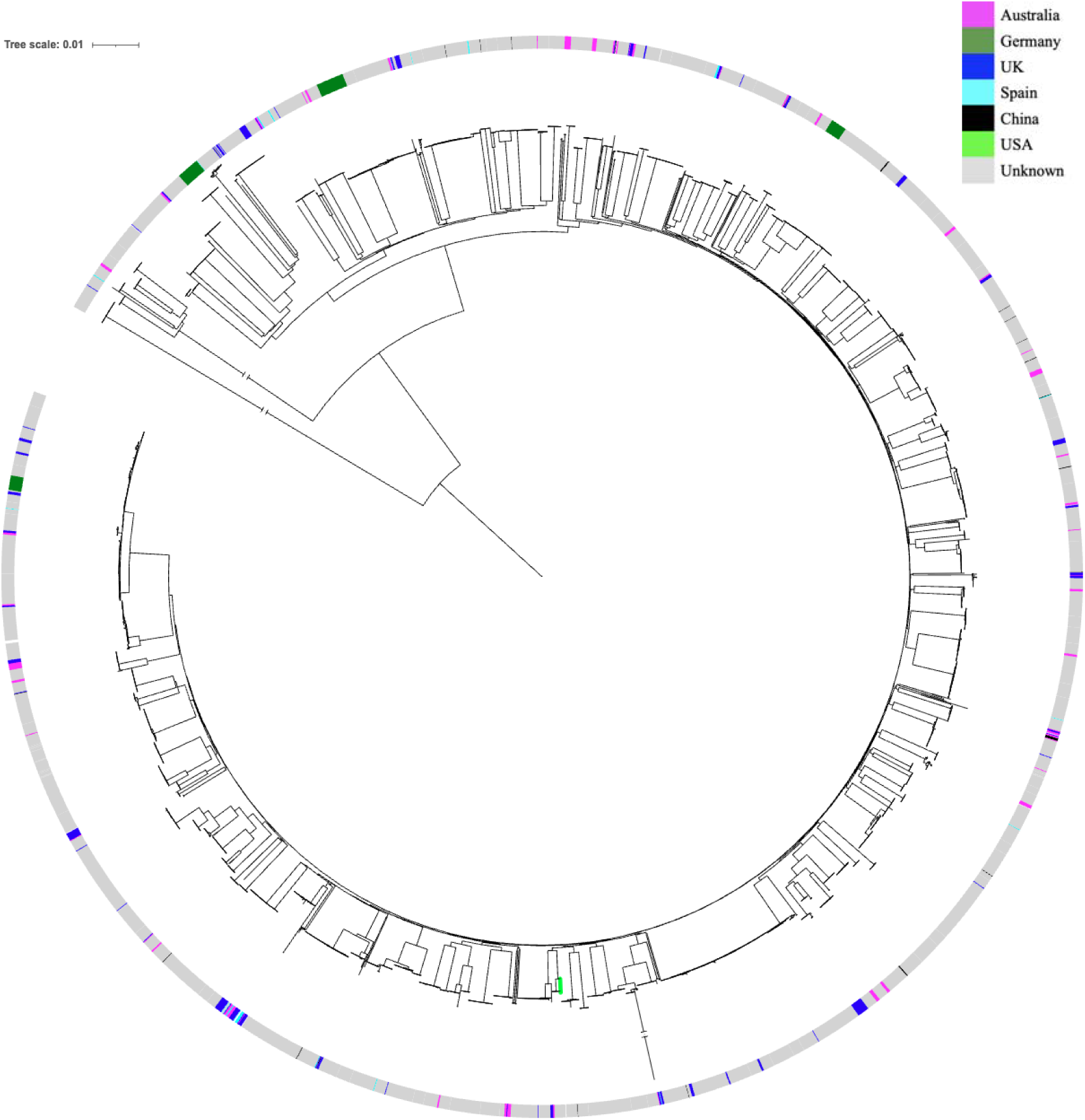
Midpoint-rooted maximum parsimony phylogenomic analysis of all publicly available bronchiectasis lung-derived *Pseudomonas aeruginosa* isolates (*n=*4575) and 137 *P. aeruginosa* from our study (total *n=*4,712). PAO1 was used as the read mapping reference. Long branch lengths were truncated to aid visualisation of clade structure, as depicted by line breaks. The bright green node depicts reference strain PAO1.

## DISCUSSION

Numerous studies have investigated AMR *P. aeruginosa* in CF, bloodstream infections, and nosocomial infections. However, research detailing the prevalence and genomic epidemiology of AMR *P. aeruginosa* in bronchiectasis is limited, with a noticeable dearth of data from Australia. To address this gap, we investigated AMR prevalence, underlying genetic mechanisms, population structure, and within-host evolutionary dynamics of *P. aeruginosa* isolates from a contemporary Australian bronchiectasis cohort.

Approximately three-quarters (75.9%) of our Australian *P. aeruginosa* isolates were resistant to at least one clinically relevant antibiotic, with 46.7% AMR, 19.7% MDR, and 9.5% XDR. Fortunately, no pan-drug resistant isolates were identified. Our MDR incidence was high compared to several prior bronchiectasis studies, including a retrospective observational UK study (11%; *n*=155 participants) (22), a recent Chinese study by Xu *et al.* (16.3%; *n=*306) (20), and a small Spanish study by Cabrera *et al.* (16%; *n*=43 isolates) (21), but slightly lower than a Chinese study by Gao *et al.* (23.1%; *n*=147 isolates) (23). Compared to the Cabrera and Xu studies, we found considerably higher AMR rates towards CIP (64.2% vs. 44.2%, 39.2%), FEP (44.5% vs. 7.0%, 10.8%), C/T (29.2% vs. 0.0%, N/A), ATM (21.2% vs. 7.0%, N/A), and MEM (15.3% vs. 9.3%, 4.6%) respectively, with our AMR towards TZP (8.8%) lower than Xu (11.8%) but higher than Cabrera (4.7%) (20,21). In contrast, we observed lower AMR rates for IPM (19.0% vs 32.6%), AMK (11.7% vs 18.6%), and TOB (10.2% vs 18.6%) compared to Cabrera *et al.*, although Xu *et al.* reported substantially lower aminoglycoside AMR (TOB 2.0%, AMK 0.3%) than our cohort.

Our high CIP resistance rate reflects its widespread use in Australia as the sole oral therapy option for exacerbations and its status as a first-line therapy recommendation according to multiple international respiratory guidelines (60–64). However, our rate was substantially higher than other bronchiectasis studies (range: 13-44%) and even Australian CF cohorts (46.0%) (15,20–23). We found that CIP resistance was driven by chromosomal *gyrA* or *gyrB* mutations rather than acquisition of an aminoglycoside modification enzyme, consistent with other bronchiectasis studies (20,65). Given the high prevalence of these AMR determinants in our local bronchiectasis population, the introduction of routine CIP AMR genotyping into clinical practice (66) would assist with antimicrobial stewardship efforts and may reduce treatment failure rates. Ultimately, broader geographic differences in AMR rates reflect variations in local antibiotic stewardship and prescribing practices, coupled with the underlying genetics of circulating *P. aeruginosa* populations. Given the low (<1%) prevalence of AMR-conferring MGEs in bronchiectasis-derived *P. aeruginosa*, our findings emphasise the importance of accurate and up-to-date AST data collection and AMR detection at local scales, and the critical need to identify chromosomally encoded AMR determinants, not just MGEs, in AMR determinant prediction pipelines.

Although comprehensive AMR surveillance data targeting Australian bronchiectasis cohorts is limited, a large study of 934 Australian CF isolates found a 35% MDR rate (15), supporting the notion of increased AMR rates in CF compared to bronchiectasis cohorts (67). Nevertheless, the AMR and MDR rates observed in our cohort remain concerningly high compared to broader national averages. Our MDR rate of 19.7% is six-fold higher than the 3.2% reported MDR rate for bloodstream infections described by the Australian Group on Antimicrobial Resistance (AGAR) 2023 report (68,69), and is markedly higher than the national average *P. aeruginosa* AMR rate (< 7% for all five antimicrobials tested) reported in the Antimicrobial Use and Resistance in Australia (AURA) 2023 report (70). Additionally, the 2023 AGAR report found a significantly higher 30-day all-cause mortality rate for MDR (46.2%) compared to non-MDR (18.2%) *P. aeruginosa* infections, raising concerns for respiratory cohorts. Our findings highlight that bronchiectasis is an important and distinct patient cohort that experiences higher contemporary AMR rates than is currently appreciated.

CST is a critically important last-line antibiotic for *P. aeruginosa* treatment, as it typically remains effective against otherwise untreatable infections due to its unique mechanism of action (71,72). Although global CST resistance remains low, rates have steadily increased from 2% in 2006–2010 to 5% in 2020–2023 (73). Most previous *P. aeruginosa* bronchiectasis studies have either not assessed CST resistance or have not identified any resistant strains (20–24), except for Xiao *et al.*, who identified a single CST resistant (*n=* 1/66) isolate in China (65). The PROMIS-I and PROMIS-II randomised control trials, which investigated CST nebuliser efficacy in bronchiectasis across 12 countries, including Australia, reported modest *P. aeruginosa* CST resistance rates (2.4% and 9.0%, respectively, in the CST group vs. 1.3% and 0.9%, respectively, in the placebo group); however, the geographic origin of these CST resistant isolates was not reported (74). Furthermore, despite its critical importance, Australia’s national registries (AURA, CF Data Registry, and the Australian Bronchiectasis Registry) do not routinely report CST resistance rates (53,70,75,76).

To the best of our knowledge, our study reports the first documented CST-resistant *P. aeruginosa* isolate (SCHI0273.S.1) isolated from an Australian bronchiectasis cohort. The isolate was recovered from an elderly participant on long-term nebulised CST with severely impaired lung function (FEV_1_: 39.5%) and a severe BSI score of 14. CST resistance is predominantly driven via activation of two-component systems, frequently PmrAB or PhoPQ, which mediates the addition of positively charged moieties to lipid A, preventing the initial electrostatic interactions of CST (71,72,77,78). We identified a missense mutation in *pmrB* (Val136Met), which was recently identified in an experimental CST evolution study (79), and a *phoQ* mutation (Gln263His), reported once prior in a hospitalised patient with a CST-resistant *P. aeruginosa* infection of unspecified origin (80). Although this novel combination of mutations is strongly suspected to confer CST resistance, functional verification is required to confirm their role.

Β-lactams are a cornerstone of *P. aeruginosa* treatment in BE cohorts, acting as the primary intravenous option in cases of oral CIP therapy failure. It is therefore unsurprising we identified numerous mutations associated with β-lactam resistance, many deleteriously impacting OprD, PBPs, and efflux pump regulators (Table 2, Table S2). Under baseline conditions, AmpR represses *ampC* transcription; however, deleterious variants can abrogate its repressor function, leading to greatly increased *ampC* expression and β-lactam hydrolysing capacity (81,82). Although *ampR* mutations are frequently described in CF isolates (58,81) they are reportedly rare in bronchiectasis cohorts, with Hilliam and colleagues reporting a 1% *ampR* mutation rate (27). Similarly, AmpC Val239Ala, identified in six (4.4%) of our bronchiectasis isolates, has been previously reported in CF cohorts, and is associated with CAZ and TZP resistance (83). Interestingly, our cohort had higher prevalences of AmpC Val239Ala (4.4% vs 1.0%, *p <*0.001) and AmpR Asp135Asn (5.1% vs 2.2%, *p=*0.029) determinants compared with the global bronchiectasis isolates, indicating a potential overrepresentation of AmpR/AmpC-driven β-lactam resistance in Australian bronchiectasis cases that deserves further exploration.

In addition to *ampC* upregulation, cell permeability alterations play an important role in conferring β-lactam resistance. As such, we found three nonsense OprD variants (Phe36fs, Trp65*, Gln79fs) that occurred between residues 1-82, affecting the β-barrel structural region (Table 3) and rendering the cell impermeable to carbapenems (84). Additional deleterious OprD AMR determinants (Gln166fs, Phe308fs, Arg342fs, Trp417*, Ala420fs) were also identified in our cohort. In all cases, isolates with these mutations were resistant to imipenem, and most were resistant to meropenem, consistent with the known role of certain OprD variants to confer carbapenem resistance (25). Similarly, other bronchiectasis studies have identified numerous OprD variants responsible for IPM resistance, rather than acquired carbapenemases, highlighting the crucial importance of OprD mutation in conferring carbapenem AMR in *P. aeruginosa* (20,21). In strains resistant to all four tested β-lactam classes (carbapenems, cephalosporins, penicillins, monobactams), multiple mutations affecting *ftsI*, *nalD*, *oprD*, and *ampR* were present; all likely contribute to this MDR phenotype (85–87).

Comparison of AMR rates towards the β-lactam/β-lactamase inhibitor combinations, TZP and C/T, identified three-fold higher C/T resistance, despite all participants being C/T treatment-naïve prior to our study. Among the TZP resistant isolates, we identified a significant association with XDR status (*p=*<0.001), which likely reflects higher TZP use to treat severe infections with recalcitrant strains (63). It is less clear why we observed high C/T resistance rates in our cohort given that this phenotype was not associated with AMR/MDR/XDR status (*p>*0.05), prior or active C/T use, or cross-resistance. One possibility is that some Australian strains possess intrinsic C/T resistance, encoded by geographically distinct yet undefined AMR resistance mechanisms. Another more likely explanation is that current EUCAST disc diffusion criteria for C/T (< 23mm for AMR strains (42)) lack sufficient stringency for AMR strains, as most of our resistant isolates measured between 20-22mm (*n=* 29/40). Further research is clearly required to identify the AMR determinants driving baseline C/T resistance, and to more effectively categorise disc diffusion criteria for this newer antibiotic.

No high-risk MGEs were identified in our cohort, such as metallo-β-lactamases (e.g. *bla*_VIM_ or *bla*_IMP_ carbapenemases), extended spectrum β-lactamases, or mobile CST resistance (*mcr)* genes. However, given the recent 2024 Australian CARAlert Annual Report found a 29.3% increase in carbapenemase-producing *P. aeruginosa* 2023 to 2024, the lack of MGEs in this cohort is likely temporary (88). Nevertheless, our current results further support our prior observation that Australian *P. aeruginosa* strains evolve AMR predominantly via chromosomal mutations, rather than acquisition of AMR-encoding MGEs (25).

Global *P. aeruginosa* epidemiology is largely driven by the dissemination of international high-risk clones. These STs are distinguished by their high MDR rates, environmental persistence, frequent hospital transmission, and successful lung colonisation (89). In our cohort, we identified five high-risk clones (Table S3), with ST274 being the most prevalent (*n=*9, 6.6%), along with several common Australian lineages, including ST155 (*n=*5/137, 3.6%), ST782/AUST-08 (*n=*2/137, 1.5%), and ST260 (*n=*1/137, 0.7%), but no high-risk Australian CF clones such as ST649/AUST-01, ST266/AUST-02, or ST801/AUST-06 (90).

The high ST274 rate in our population is concerning as this ST is also common in people with CF, where it is associated with high AMR rates (89–92). Xu *et al.* also found ST274 (*n=* 16/306, 5.2%) to be the most common ST in bronchiectasis (20). ST274 has been reported previously in Germany and two UK studies; however, these bronchiectasis studies found greater proportions of other international clones that were either absent (ST17 [Clone C]), or less prevalent in our study (ST253 [*n=*7/137, 5.1%], and ST395 [*n=*1/137, 0.7%]) (27,93,94). ST395 has not been reported in Australian CF populations, only bronchiectasis cohorts; therefore, a potential association between ST395 and bronchiectasis may exist, although this requires further validation.

On a phylogenomic scale, we found no distinct geographic clustering when comparing our Australian isolates to the global dataset (Figure 5). This panmictic population structure has been noted in prior studies investigating both clinical and environmental *P. aeruginosa* from similar geographical regions (25,95), and other bronchiectasis studies (20,27,65), demonstrating that *P. aeruginosa* clones are highly transmissible across geographic boundaries.

In people with bronchiectasis, *P. aeruginosa* lung infections can persist for years, even decades, despite aggressive antimicrobial therapy (27). Our phylogenomic analysis confirmed persistence (i.e. infection with the same strain for ≥6 months) in 13 participants, with the longest example being at least 9 years (Table S3). In two cases, we observed substantially higher mutation rates; specifically, in P25 this elevated rate was attributed to a 50kb recombination event with an unsampled *P. aeruginosa* strain, resulting in >2,000 SNP differences over just a six-month period (Figure 4). In P09, latter strains accumulated ∼1,000 mutations over an 8.7-year period (SNP accumulation rate of 19.0 substitutions per million base pairs/year), noticeably higher than the average 1.16 substitutions per million base pairs/year for the remaining 11 participants (Figure 4). No high-consequence MutL, MutS, or UvrD variants were identified in P09 isolates, with the latter strains encoding MutS Asp552Asn; however, we deem this variant as speculative given that no other MutS missense variants (e.g. Gly31Ser, Ala187Val, Ile579Val) were definitively associated with hypermutation. In contrast, 9/59 (15.3%) of participants (P04, P30, P31, P40, P41, P46, P47, P49, P56) encoded high-consequence MutS or MutL variants, indicating that these participants harboured hypermutable *P. aeruginosa* (Table S3). Hypermutation and recombination thus represent key adaptative strategies adopted by *P. aeruginosa* in bronchiectasis airways to increase genetic heterogeneity, in turn accelerating the transition towards chronic persistence and potential AMR acquisition.

Alongside hypermutation, we observed several convergent within-host evolutionary signatures in persistent isolates, consistent with the transition to a chronic, sessile, biofilm-driven lifestyle (Table S3, Table S4). Deleteriously affected loci comprised those associated with motility/chemotaxis (*aer*, *chpA*, *chpE*, *cupA2-5*, *cupB3*/*5*/*6*, *cupC1*/*2*, *fleS*, *fleR*, *flgK*, *flgL*, *flhA*, *fliC*, *fliI*, *pilA*, *pilB*, *pilD*, *pilG*, *pilJ*, *pilM*, *pilN*, *pilP*, *pilQ*, *pilV*, *pilW*) lipopolysaccharide synthesis or remodelling (*arnD*, *arnT*, *gmd*, *galU*, *lpxO2*, *migA*, *pagL*, *rmlA*, *rmd*, *waaC*, *waaL*, *wapR*, *wbpM*, *wbpY*, *wbpZ*) loci, virulence and quorum sensing (*exsA*, *exoS*, *exoT*, *exoY*, *lasA*, *lasI*, *lasR*, *mvfR*, *pscB*, *pscP*, *pqsA*, *pqsB*, *vreA*/*I*/*R*, *xcpR*), iron and haem acquisition (*fiuA*, *fpvB*, *fpvI*, *fpvR*, *hemO*, *hasD*, *hasR*, *pchD*, *pchE*, *pchF*, *tonB2*), and biofilm exopolysaccharide/mucoidy production (*algG*, *algL*, *algU*, *mucA*, *mucD*, *pelB*, *pelD*, *pelE*, *pelG*, *pslH*, *pslL*, *rpoS*, *tpbB*, *vfr*, *wspR*, *yfiR*) (Table S3, Table S4). These hallmark adaptations are consistent with the switch to chronic persistence in the hostile airway environment previously reported in *P. aeruginosa* and other bacterial respiratory pathogens (96). Such mutations provide highly favourable conditions for *P. aeruginosa* by enhancing immune evasion (especially via reduced immunogenicity), rewiring metabolic pathways to reduce energy-expensive cellular processes, and passively increasing antimicrobial and bacteriophage resistance via the production of highly protective, mucoid-rich biofilms and altered or absent lipopolysaccharide moieties. In addition, we identified 28 strains encoding the PA5438 (GavR) Ser275_Arg277del variant, which is strongly associated with the transition from acute to chronic *P. aeruginosa* infection (32). Taken together, these substantial genomic adaptations drive a high rate of chronic persistence that closely mirrors CF cohorts, ultimately establishing a formidable barrier to successful immunological and antimicrobial eradication.

Our study has several limitations. First, despite our isolates being collected from multiple tertiary centres, they were collected from the same subtropical geographical region (Southeast Queensland, Australia) and are therefore not fully representative of clones found across Australia. Secondly, sampling was opportunistic, with longitudinal sampling only available for a subset of participants, leaving gaps in our understanding of parallel within-host evolution pathways, transient infections with different lineages, and additional transmission and population dynamics. Third, functional validation of novel AMR determinants, including those encoding CST or C/T resistance, was outside of scope for this work, but represent an important avenue for future research given the importance of these antibiotics in contemporary *P. aeruginosa* treatment regimens.

## CONCLUSION

Our findings shed important light on contemporary AMR prevalence, genetic diversity, and within-host dynamics of *P. aeruginosa* from an Australian bronchiectasis cohort. We found higher than anticipated rates of AMR/MDR/XDR, including the first reported CST resistant isolate in our region. In all cases, AMR was conferred by chromosomal mutation rather than MGE acquisition. The detection of persistent infections, within-host adaptive evolution, and multi-lineage co-infections further highlights the dynamic and heterogeneous nature of *P. aeruginosa* in the bronchiectatic airway. Our findings emphasise the need for accurate AST and genomic sequencing of *P. aeruginosa* from bronchiectasis airways to assist with the development of personalised treatment strategies. Such efforts are needed to improve long-term outcomes for people living with bronchiectasis.

## Data Availability

All data included in the study is publicly available in BioProject PRJNA1330417 and PRJNA1325058

## ACKNOWLEDGEMENTS

We thank Barry Pritchard and Claire Winterbourne (Sunshine Coast Health Institute) for laboratory management support, The Mater Research David Serisier Respiratory Biobank curators (Megan Martin, Lucy Burr, and Simon Bowler) for providing access to retrospective bronchiectasis sputa, and the Queensland Cyber Infrastructure Foundation Ltd at University of Queensland for Bunya high-performance computing support.

